# A qualitative exploration of barriers to efficient and effective Structured Medication Reviews in Primary Care: Findings from the DynAIRx study

**DOI:** 10.1101/2024.04.18.24303004

**Authors:** Aseel S Abuzour, Samantha A Wilson, Alan A Woodall, Frances S Mair, Andrew Clegg, Eduard Shantsila, Mark Gabbay, Michael Abaho, Asra Aslam, Danushka Bollegala, Harriet Cant, Alan Griffiths, Layik Hama, Gary Leeming, Emma Lo, Simon Maskell, Maurice O’Connell, Olusegun Popoola, Sam Relton, Roy A Ruddle, Pieta Schofield, Matthew Sperrin, Tjeerd Van Staa, Iain Buchan, Lauren E Walker

## Abstract

**Introduction:** Structured medication reviews (SMRs), introduced in the United Kingdom (UK) in 2020, aim to enhance shared decision-making in medication optimisation, particularly for patients with multimorbidity and polypharmacy. Despite its potential, there is limited empirical evidence on the implementation of SMRs, and the challenges faced in the process. This study is part of a larger DynAIRx (Artificial Intelligence for dynamic prescribing optimisation and care integration in multimorbidity) project which aims to introduce Artificial Intelligence (AI) to SMRs and develop machine learning models and visualisation tools for patients with multimorbidity. Here, we explore how SMRs are currently undertaken and what barriers are experienced by those involved in them.

**Methods:** Qualitative focus groups and semi-structured interviews took place between 2022-2023. Nine focus groups were conducted with doctors, pharmacists and clinical pharmacologists (n=21), and three patient focus groups with patients at high-risk of rapidly worsening health from multimorbidity (n=13). Five semi-structured interviews were held with 2 pharmacists, 1 trainee doctor, 1 policy-maker and 1 psychiatrist. Transcripts were analysed using a thematic approach.

**Results:** Two key themes limiting the effectiveness of SMRs in clinical practice were identified: ‘Medication Reviews in Practice’ and ‘Medication-related Challenges’. Participants noted limitations to the efficient and effectiveness of SMRs in practice including the scarcity of digital tools for identifying and prioritising patients for SMRs; organisational and patient-related challenges in inviting patients for SMRs and ensuring they attend; the time-intensive nature of SMRs, the need for multiple appointments and shared decision-making; the impact of the healthcare context on SMR delivery; poor communication and data sharing issues between primary and secondary care; difficulties in managing mental health medications and specific challenges associated with anticholinergic medication.

**Conclusion:** SMRs are complex, time consuming and medication optimisation may require multiple follow-up appointments to enable a comprehensive review. There is a need for a prescribing support system to identify, prioritise and reduce the time needed to understand the patient journey when dealing with large volumes of disparate clinical information in electronic health records. However, monitoring the effects of medication optimisation changes with a feedback loop can be challenging to establish and maintain using current electronic health record systems.

## Introduction

Structured medication reviews (SMRs) were introduced in the United Kingdom (UK) in October 2020 and incorporated into the NHS England Directed Enhanced Service contract (DES) for 2021.(1) SMRs represent a National Institute for Health and Care Excellence (NICE)-approved clinical intervention facilitating shared-decision making between clinicians and patients, to inform treatment decisions. The objective is to reduce medication-related harm in patients with complex or problematic polypharmacy. While General Practitioners (GPs), pharmacists and advanced nurse practitioners (ANPs) who meet training criteria can conduct SMRs, the prevailing expectation is for clinical pharmacists within Primary Care Networks (PCNs) to assume primary responsibility as a commissioned service.(2) The varied methods employed by PCNs to proactively identify patients suitable for SMRs, and conduct these reviews, is contingent on available resources and capacity. Anecdotal evidence suggests that PCNs currently use limited digital tools, such as searching electronic health records (EHR) based on the total number of drugs prescribed or disease codes, to identify patients at risk of medication-related harm.

There is sparse empirical evidence reporting on the implementation of SMRs, their impact on patient outcomes, and the challenges faced by healthcare professionals (HCPs) and patients during SMRs.(3, 4) This scarcity of evidence is unsurprising given that SMRs were introduced in 2020 amidst the COVID-19 pandemic. Nonetheless, estimates suggest a percentage reduction in per-patient medicines following an SMR ranging from 2.7% to 9.9%, with up to 19.5% reduction in use for the highest-risk group in care homes.(5)

Patients with complex multimorbidity and polypharmacy, whose medicines have not been optimised are at risk of adverse outcomes and medication-related harm.(6) The use of data from EHRs to develop evidence-based digital health tools can be a promising resource to assist HCPs in conducting targeted, efficient and effective SMRs. The NIHR-funded DynAIRx study (Artificial Intelligence for dynamic prescribing optimisation and care integration in multimorbidity) aims to develop AI-driven tools that integrate information from electronic health and social care records, clinical guidelines and risk-prediction models.(7) The DynAIRx project will produce machine learning models, dashboards, and different tools including Causal Inferencing to provide clinicians and patients with evidence-based information to prioritise patients at most risk of harm and/or patients most likely to benefit from SMRs. Aligned with the NICE multimorbidity guidelines, DynAIRx will focus on three patient groups at high-risk of rapidly worsening health from multimorbidity: (a) individuals with mental and physical health co-morbidity, in whom the prescribing for mental health improvement can lead to adverse physical health consequences; (b) those with complex multimorbidity (four or more long-term health conditions taking ten or more drugs); and (c) older people with frailty who are at high risk of adverse outcomes.

The initial step towards introducing AI-driven prescribing support tools into clinical practice involves understanding the current scope of work, how SMRs are presently undertaken and by whom, the time required in real-world clinical practice to undertake them, and crucially, the barriers to effective SMR implementation. The aim of this study was to explore how SMRs are undertaken and what barriers those undertaking them (and receiving them) experience.

## Methods

### Participants and recruitment

This study sought to recruit health care or management professionals working in health care settings (primary care in the community or secondary care in hospital services) across the UK where review of prescription medications is a regular part of the clinical workload. This included those working in General Practice, secondary care hospital services (geriatric medicine, clinical pharmacology, falls clinics, mental health practitioners), clinical commissioning of services or management of clinical services (practice managers), and pharmacists, including PCN pharmacists (those involved in conducting SMRs across several neighbouring GP practices). Patient and carer representatives of the three key multimorbidity groups outlined above were also invited, patients with (a) multiple and physical co-morbidities; (b) complex multimorbidity; (c) older people with frailty. This included recruiting adult individuals (over the age of 18) with/or caring for someone with multiple (4 or more) long-term health conditions, co-existing mental and physical health problems, prescribed ≥10 regular medications, frailty.

Purposive sampling identified potential HCP participants that were known to be involved in medicines optimisation services through the researchers own clinical and professional networks. Snowballing (wherein research participants were asked to assist the recruitment by attempting to identify other potential participants) was employed to identify contacts through existing service providers along with advertisement in GP forums and at national events for individuals participating in clinical polypharmacy research.(8) Purposive sampling of potential patient representatives were identified through advertisement across the NIHR Applied Research Collaboration public advisor networks and through research databases at the researchers host institutions. Potential participants were provided with study information and an invitation to participate. Participants received comprehensive briefings from researchers about the study, and written consent was obtained prior to the focus group or interview participation. Withdrawal of consent was permitted at any stage, even after the focus group or interview.

### Ethical approval

The Newcastle North Tyneside Research Ethics Committee (REC reference:22/NE/0088) granted ethical approval for the DynAIRx study.

### Data collection

Data collection occurred from November 2022 to November 2023. Focus groups and semi-structured interviews were conducted to gather participant views. Focus group topic guides and interview schedules were developed and refined by the clinical members of the research team (LW, AA, AW, FM, AG) and tailored to HCP and patient groups. The topic guides (see S1 Appendix) included questions exploring the experience of conducting or receiving SMRs, barriers to undertaking them and opinions on key medication challenges in multimorbidity groups from both the clinician and patient perspective. Sessions occurred in person and online (via Microsoft Teams), lasting from 49 to 109 minutes. Audio recordings underwent verbatim transcription and anonymisation to remove any potentially identifiable information. Each participant was assigned a code, and recordings were subsequently deleted. Data collection and analysis occurred concurrently.

### Data analysis

Transcripts were imported into QSR NVivo 12® and analysed using thematic analysis.(9) Transcripts were read to familiarise researchers with the data. Inductive reasoning guided the initial coding by AA and SW, who collated and examined codes to identify themes. The coding team (AA, SW, LW, AW, FM) reviewed and discussed initial themes. The remaining dataset underwent hybrid inductive and deductive thematic analysis, with codes and themes iteratively revised. Themes were defined and supported by quotes, maintaining detailed notes to ensure analytical rigour and plausibility.(10)

## Results

Nine focus groups with HCPs (n=21) and 3 patient focus groups (n=13) were conducted. A further five semi-structured interviews with HCPs took place (see Table 1 for details).

**Table 1.**
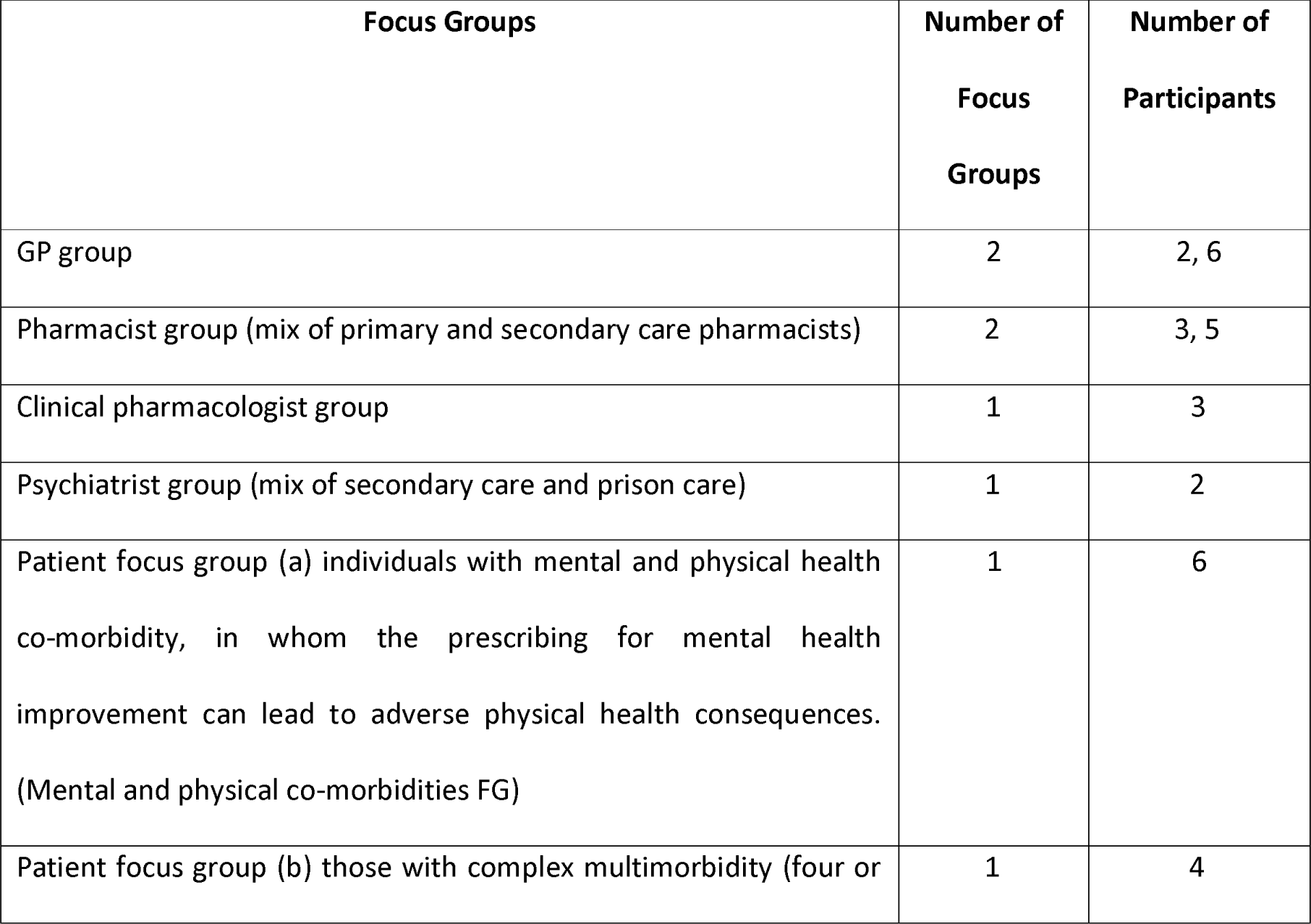
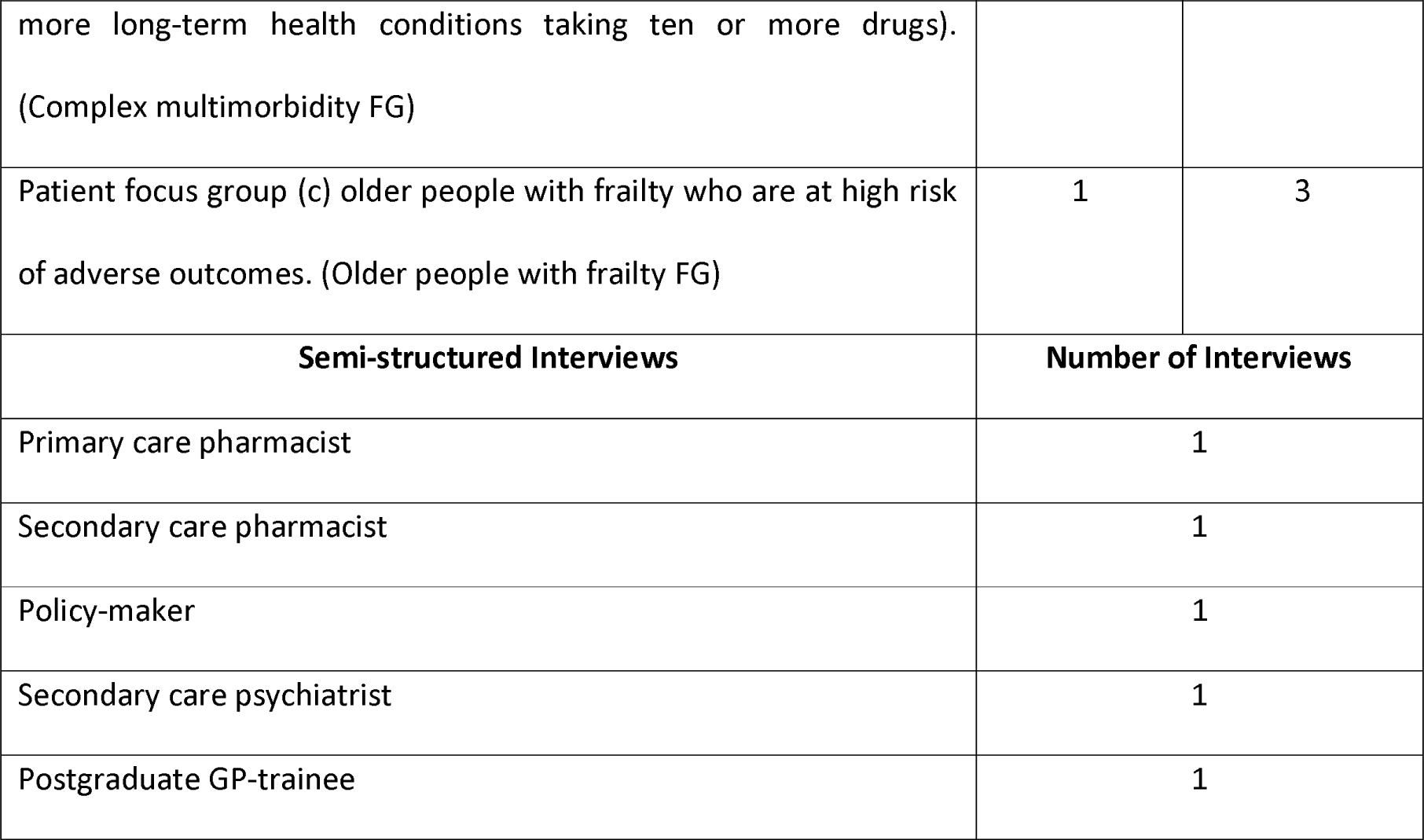
Table showing participant type and number of participants that took care in the focus groups and semi-structured interviews.

Two overarching themes emerged from analysis of the HCP and patient interviews and focus groups, within which a number of sub-themes emerged:

1. Medication reviews in practice

a. *Limited availability of digital tools to assist in identifying and prioritising patients for a SMR*
b. *Organisational challenges and patient factors affecting patient engagement for a SMR*
c. *Time consuming “detective work”*
d. *SMRs require multiple appointments*
e. *Influence of healthcare context on delivering SMR*
2. Medication-related challenges

a. *Poor communication and data sharing between primary and secondary care*
b. *Difficulties managing mental health medication for prescriber and patient*
c. Challenges around anticholinergic medication optimisation for prescriber

**Fig 1.**
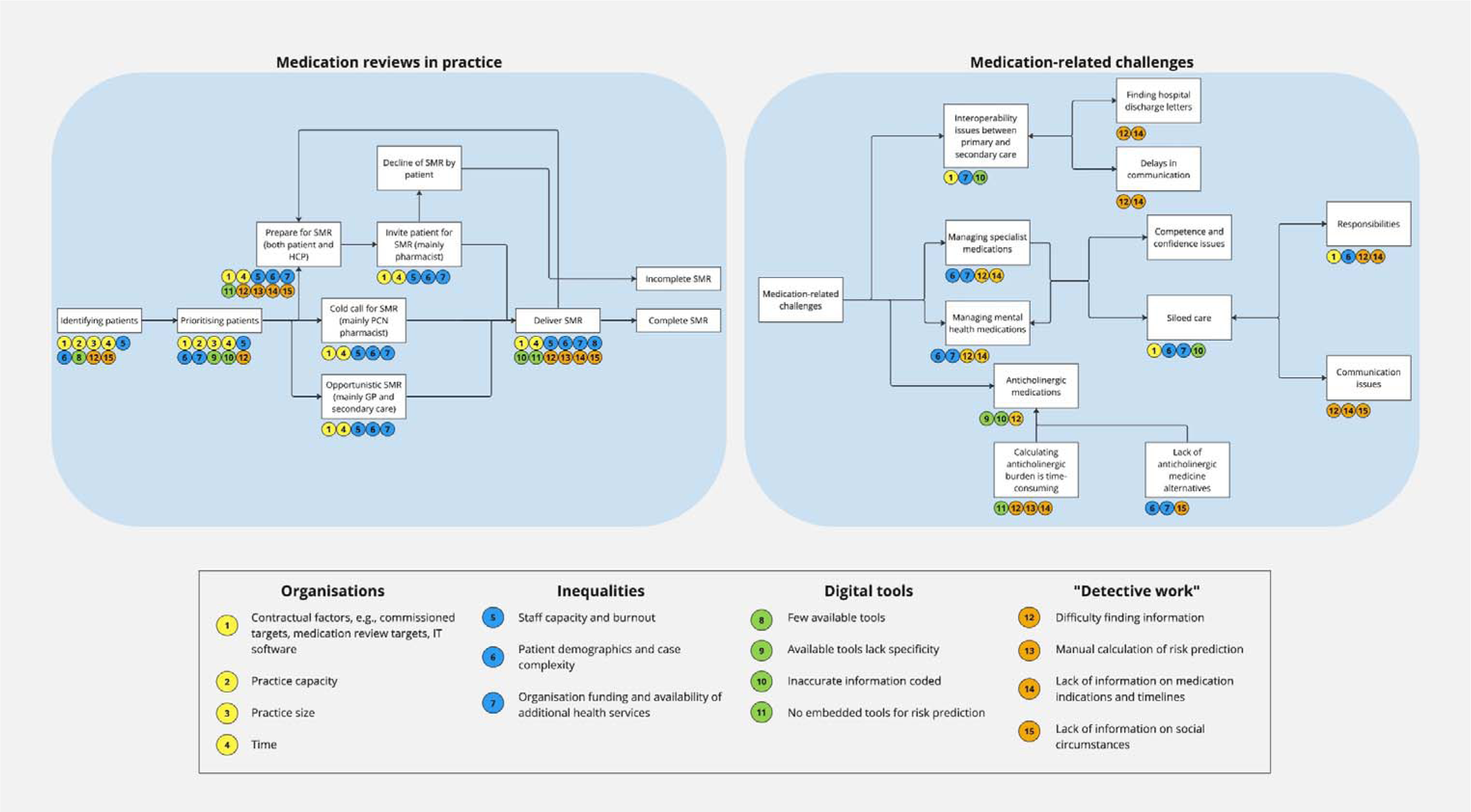
displays each key theme from this study and a detailed list of the barriers to each stage of the SMR process.

### 1. Medication reviews in practice

#### a. Limited availability of digital tools to assist in identifying and prioritising patients for a SMR

PCN pharmacists reported proactively identifying and prioritising patients to conduct SMRs. Patient identification was based on the criteria outlined by the DES GP contract, which includes patients in care homes (both nursing and residential facilities), individuals with complex multimorbidity and polypharmacy, urgently referred patients, older individuals encountering medication-related harms, and patients recently discharged from hospital. HCPs also referenced using available healthcare management automated search tools within the EHR, including ‘Ardens search’ (11) and ‘ProActive Register Management (PARM) diabetes’ (12), to identify pre-determined conditions, prescribing alerts and other variables that form part of the practice workload. They also used targets from the Investment and Impact Fund (IIF) for patient identification (IIF is an incentive scheme where PCNs can improve health and the quality of care for patients with multimorbidity), which participants described as beneficial but a waste of resources in the context of SMRs which should focus on patients with complex multimorbidity and polypharmacy.(13)

Since the introduction of EHRs in the NHS, HCPs are expected to assign ‘SNOMED codes’ to document patients with specific diagnostic, symptom or treatment codes in a logical hierarchical manner to specify clinical information.(14) These codes facilitate searches for specific medical conditions, symptoms and treatments within the GP EHR, facilitating the identification of individuals requiring an SMR. Pharmacists noted that EHR searches do not adequately consider the complexity of patients, making it challenging to stratify those that would benefit most from an SMR. Additionally, participants highlighted a lack of sensitivity and specificity in the current search mechanisms (meaning the searches either do not identify all the intended patients or identify too many).

> “*The actual indicator that my team has been focusing on is supposed to be the ones where patients are prone to medication errors … but when I actually look at the patients, I haven’t got a clue why the actual computer system has decided that most of the time.”* (Participant 1, Pharmacist FG1)

HCPs were concerned that the identification of patients who would benefit from an SMR could exceed the clinical capacity of the staff available to meet the need. They felt that any digital tool used to prioritise patients’ needs should match the clinical capacity of the practice.

> “*The tools have to be a bit cleverer … We can generate a list of patients today … PCNs at the moment essentially do that, but what you have to do is almost the list that’s generated to the capacity … People would not switch it on if they felt that it could generate lots of patients you would not then see.”* (Policy-maker interview)

#### b. Organisational challenges and patient factors affecting patient engagement for a SMR

GPs or secondary care clinicians (excluding clinical pharmacologists) often conducted opportunistic medication reviews, compared to the proactive SMRs conducted by PCN pharmacists. In alignment with the DES requirements, HCPs described how the task of conducting an SMR was contingent upon organisational contracts, practice size, and staff availability. The presence of a PCN pharmacist for SMRs facilitated streamlined tasks, enabling GPs to focus on patients with more complex medical profiles. GPs voiced concerns about burnout in areas where demand for SMR exceeded the clinical capacity to undertake them. This challenge was particularly pronounced in regions of lower socioeconomic status where patients often presented with complex multmorbidity and polypharmacy at a younger age, especially those with co-morbid mental and physical health problems. Moreover, respondents felt that patients residing in deprived areas were less likely to attend scheduled SMRs, compelling GPs to resort to opportunistic reviews. This highlights potential inequity in access to SMRs and overall health surveillance.

> *“In the poorer area of the practice there’s no clinical pharmacist, that’s all done opportunistically, if done at all, by the GP partners there. I think that there’s a couple that are approaching burn out, if not complete burn out and the practice is almost run by locums. So, when I’m going in there, it’s quite tough and I will often see medications that are inappropriately prescribed, polypharmacy, several of the same drugs, and I will opportunistically undertake a structured medical review.”* (Participant 3, GP FG1)

#### c. Time consuming “detective work”

Whether HCPs identified patients proactively or opportunistically, the preparation time for a medication review ranged from 10 minutes to 1 hour. Several factors influenced this preparation time, including the availability of information, case complexity, barriers to accessing information, information density, and time constraints. The challenge in finding and collating information within the patient’s clinical records constituted a significant portion of the preparation time. For instance, discharge letters from hospitals are often located as attachments within the patient record, requiring HCPs to locate and read the letter. These necessary preparatory activities take away from the face-to-face time available with the patient.

> “*Probably double the amount of prep time than it was actually with the patient. I mean, granted we did spend a while with the patient because we both like to talk, and the patient certainly did, but I think, and that’s the problem, isn’t it? You get the best information out of your patient when you let them talk and you let them tell you lots of things that you wouldn’t normally ask, but you haven’t got the time to do that so it’s tricky isn’t it to find the balance. But the biggest thing with the prep time was getting the information.”* (Participant 3, Pharmacist FG1)

HCPs also conveyed frustration regarding the substantial time required to determine the original indication for a particular prescription and the ongoing necessity for it, even during major transition periods such as a patient’s admission to a care home.

> “*We don’t get enough actual structured reviews, so they’ll be getting put on medication, people in care homes, and then left on those medicines. There’s no recognition of the changes. As you move in a care home, you’re generally more frailer, your renal function, haematic function might not be as great and, you know, you’re not moving as much, so your need for some medicines might not be as great as it was when the medicine was first started.*” (Policy-maker interview)

Although it is possible within the EHR system to link the prescription of an individual drug to its clinical indication, in clinical practice this is usually not done as it is time consuming. As such, indications for prescribed medicines are recorded in the free text for the consultation which can easily become obscured over time within the extensive information contained in the clinical record. Examining the clinical free text for this information was emphasised as a challenge in efficiently conducting SMRs.

> “*Although in my letters I would clearly state to the GP why I am prescribing the second line antipsychotic just so that people know, but over time that tends to get lost, the rationale for that prescribing tends to get lost and before you know you leave post, somebody else comes and begins to increase that second antipsychotic you know, so that becomes a problem.*” (Participant 1, Psychiatrist FG)

Moreover, existing EHRs are not adept at presenting patient histories in a manner conducive to HCPs pinpointing areas for potential deprescribing. This deficiency in the system leads to a cumulative high pill burden for patients, as illustrated in the quote below.

> “*At the age of [18-20], I was diagnosed with bipolar. I am now [71-74] and I have lived for that period of time on medication, a lot of medication actually … I counted the number of tablets and my boxes on my bedside the other day and there was 13 different tablets, so that is what I am being prescribed by my GP.*” (Patient 1, Mental and physical co-morbidities FG)

Patients also expressed uncertainty about the initial reasons for starting medications. Patients reported receiving medications for many years and being unsure whether the medication was still necessary.

> “*She is also on a daily injection of adult growth hormone which another consultant put her on at the time and she has been having them for probably 10 to 15 years, and no-one seems to know now who initially prescribed it and who is in charge of that. I am concerned, does she really need them? She is having them every day … Initially it was an asset to go with the immunodeficiency but now I don’t really know.*” (Patient 5, Mental and physical co-morbidities FG)

#### d. SMRs require multiple appointments

SMRs typically lasted a minimum of 30 minutes, often extending beyond this duration. The variability in duration was contingent upon the patient’s complexity and the focused nature of the review. Allowing adequate time to address broader health concerns was deemed crucial, enabling the identification of potential issues requiring deeper exploration by the clinician.

HCPs acknowledged that SMRs were not a singular event, and patients might necessitate multiple appointments for a comprehensive review. Consequently, EHR systems were recognised as needing functionality to alert HCPs to schedule additional appointments after the initial SMR, emphasising the iterative and ongoing nature of medication reviews.

> “*The first time I see patients, you want almost a bit of a holistic conversation, but actually when you start making interventions you go with what matters most to the patient or where the biggest risk is and you then table the others … You can imagine that being 2 or 3 hours in 4 different appointments before you get to the bottom of where you want to be … I think we had to contact on average about 2 to 3 times per patient, but there were more complex patients as well … I don’t think you can stop medicines or optimise medicines without seeing that patient again as least once.*” (Policy-maker interview)

Patients expressed a desire to be involved in the decision-making during reviews and valued the opportunity to discuss issues such as how medications fit into their routines and other resources that may be available to them.

> *"I’ve got a series of chronic things, take a load of pills and they’re each for separate things, and I have been concerned for years whether there’s any interaction with them, between them. And also they make me feel tired all the time and perhaps there are some of them where I could actually get rid of them."* (Patient 1, Older people with frailty FG)

Patients also emphasised the importance of their views being considered throughout the process.

> “*Success for me is when my doctor or pharmacy, I have an appointment with them, they listen to what I am saying, I feel valued.*” (Patient 1, Complex multimorbidity FG)

#### e. Influence of healthcare context on delivering SMR

The duration of conducting a SMR was also contingent on the specific healthcare setting in which it took place. One pharmacist highlighted that SMRs conducted in care homes lacked a strict time limit and were oriented towards achieving specific outcomes, such as the number of medicines deprescribed in particular patient groups. This reflected the contextual variability in the conduct and objectives of SMRs, emphasising the need for flexibility in the approach based on the healthcare environment and patient population.

> “*We were just told take whatever time you need but because we were not measured on the quantity, we were measured on the quality, and we were recorded the number of medicines basically stopped and in particular groups actually so, and then that would have gone on the report because that was the way of showing what we were doing and the basic value for money I guess*.” (Participant 1, Pharmacist FG1)

The emphasis placed on a medication list varied depending on the reviewer and the healthcare setting. A pharmacist working in secondary care articulated a tendency to allocate less attention to certain medicines in a hospital setting, prioritising focus on medications more likely to cause harm. This perspective highlights the nuanced approach that different HCPs may adopt based on their expertise and the specific context in which they operate.

> “*At the moment, the bisphosphonate would be something that I’m less concerned about it a very high acuity environment, that’s the thing that I’m probably going to, maybe if I get a chance, write in the discharge summary for the GP to check up on whether or not that’s still appropriate. Whereas I’m chasing those big harms*.” (Participant 2, Pharmacist FG1)

#### f. Factors influencing deprescribing discussions

Discussions around deprescribing between HCPs and patients were reported to be influenced by several factors. These included the specific type of medicine to be deprescribed, the patient’s willingness to discontinue the medication, sociodemographic location, availability of additional health services in the area, whether the medication was initially prescribed in primary or secondary care and the existence of pre-established stopping criteria for certain medications (e.g., bisphosphonates for more than 5 years). Additionally, HCPs and patients acknowledged a degree of reluctance to engage in deprescribing due to perceived potential risks associated with the cessation of certain medications. These multifaceted factors contribute to the complexity and individualised nature of deprescribing discussions within the healthcare context.

> “I *find it really, really difficult because all of the guidelines will say, oh, you should have this patient on statins, etc., and you think I really probably shouldn’t they’re 95, but having stopped them in the past then a patient unfortunately ends up with a stroke, they go to hospital, the hospital tells them it’s because their GP stopped their statin and puts them back on*.” (Participant 2, GP FG2)

> “*The antidepressant one is interesting. So, I did work for 9 years in a really deprived area… SSRIs for a long time and they were pretty reluctant to come off it but usually willing to accept if it didn’t work out just put them back on it. It just meant in a much more middle-class area there’s the opposite where they’re all desperate to come off it and probably coming off it far too soon. So, I don’t think it needs to be doctor-led, it seems to be more about their external pressures and there’s been a lot of areas done in deprived communities with link workers and social prescribers and I think if you’re going to look at polypharmacy in these sort of groups then that’s probably the way to do it stopping all their meds.*” (Participant 4, GP FG1)

In one case, a pharmacist highlighted the challenges associated with decision-making when optimising medicines for complex, younger patients, emphasising the impact of side-effects on their quality of life. Equally, the importance of considering quality of life in frail older adults with polypharmacy was acknowledged, although perceived as less complicated than in younger adults.

> *"I know well that’s it isn’t it, it’s not necessarily that they’re a complex medicine it’s that there is evidence to say that this can prolong your life but it’s causing them that much upset, so it’s, for me it’s not necessarily the, I can’t think of any particular group of drug its more the younger you get, you know 50 is very young and that you have got a lot of life left to live so that’s when it becomes more of a clinical decision for me that as a pharmacist I don’t feel like I would be able to make" (Participant 1, Pharmacist FG1)*

### 2. Medication-related Challenges

Potential for medication-related harm identified by our key stakeholder groups included issues with specific medicines, conditions, and risky medication combinations; mental health medications; prescriptions from specialists; anticholinergic medicines; difficulties in determining prescription timelines to assist in decision-making; challenges with younger complex patients; and siloed care.

#### a. Poor communication & data sharing between primary and secondary care

HCPs identified the challenge of extracting information from hospital discharge letters as a key source of frustration. Patients, in particular, assumed that EHRs seamlessly connected primary and secondary care, and in some cases, their community pharmacy. This assumption left patients bewildered and, in certain cases, reliant on the HCPs knowing the complete narrative behind their health records. The disjointed communication and misconceptions surrounding record integration emphasised the need for improved interoperability to enhance the continuum of care.

> “*We will be waiting a week for an outpatient letter to come through and it’s really confusing, stop this, change that, increase the dose here, and you’re kind of stuck in the middle. So sometimes the patient will have left that meeting there and it will be ‘like go and see your GP and they will do this bit’, well [that’s] not happening until I’ve got that letter. There is that real mismatch of communication.*” (Participant 1, GP FG2)

> “*When the repeat prescription came through, despite the fact that the surgery had received the discharge letter, everything was all wrong, and this is just one of those things that happens. So, you get a review and it is whether that data from that review and the story behind it and who it goes back to and whether it is acted on, I think that is important*.” (Patient 3, Complex multimorbidity FG)

Communication gaps between GPs, specialist clinicians, and patients were evident due to varying expectations. GPs expressed challenges in managing specialist medications with patients, including concerns about patients’ ability to self-manage their medicines. These challenges highlighted aspects of fragmented care between primary care and specialist clinicians. HCPs also cited difficulties and reluctance in communicating and potentially engaging in conflict with specialist doctors. Participants described specialist doctors as lacking a holistic view when prescribing for patients, favouring certain medicines, and having limited knowledge in drug interactions.

Participants noted that central nervous system medicines had complicated medication regimens and hence required more coordinated care and responsibility between the specialist prescriber and GP. Addressing these challenges calls for enhanced collaboration, knowledge exchange, and a holistic approach to patient care between primary care and specialist clinicians.

> “*I find it, with the pain management clinic, they stop medication, give you a list of all these other tablets you need to start to see how things go and then sort of leave you to it, discharge the patient in your hands and expect you to sort of manage it all. And the same thing is with migraine and headaches from neurology. That’s just a minefield … I think when you’re in specialty, you feel that you can give any sort of long protracted complicated regime and the patient is just going to manage it because that’s the only medication that you think that they’re on. So yeah, they can be quite difficult*.” (Participant 5, GP FG1)

> “*He [GP] says that we can’t actually change any medication to do with your bipolar, that has got to be done by your psychiatrists … I don’t think they would change anything to do with psychiatry*.” (Patient 1, Mental and physical co-morbidities FG)

#### b. Difficulties managing mental health medication

Mental health medication and management emerged as a consistent sub-theme across key HCP stakeholder groups, irrespective of their professional background. Both doctors and pharmacists described difficulties in monitoring and adjusting psychiatric medicines, including uncertainties about how to address specific issues related to psychiatric medicines. Participants expressed a sense of being ‘out of their depth’, particularly concerning antipsychotic medicines. They conveyed a lack of confidence in assessing the risks and benefits of antipsychotic prescribing, feeling deskilled in this specific area of medication management, and finding it challenging to safely challenge prescribers. This sense of unease prompted participants to seek ways of contacting the mental health team, only to encounter additional hurdles, such as difficulties in locating relevant information within patient records to facilitate multidisciplinary coordinated patient care.

> “*But the other one is someone with very complex psychiatric problems, still maybe under the mental health team, and I haven’t got really access to the details apart from maybe I’ve got, you know, some of the other diagnoses. But if I think maybe one of those drugs is potentially causing more harm than could then I’m not clear how then to action that and who to speak to and who were they actually seeing*” (Clinical Pharmacologist 1, Clinical Pharmacologist FG)

For example, a clinical pharmacologist explained that evaluating the success of managing antipsychotics is not as straightforward as assessing physical health conditions. This complexity may contribute to the observed lack of confidence among HCPs when it comes to deprescribing psychiatric medicines. The nuanced nature of mental health outcomes, compared to more tangible markers of success in physical health, adds an additional layer of intricacy to the decision-making process in psychiatry. This includes the complexity of managing mental health medication.

> “*The biggest challenge group that I think we face in a deprived area is the patients who are on long term opioid medication, long term neuropathic meds, they’ve probably got a diagnosis of fibromyalgia, they’ve probably got personality disorder plus / minus mental health problems. And the issues that we have is that they’ve almost been sequentially added medication on because GPs don’t really often know what to do with them unless you have a special interest in that field like I do. And when they go and see pharmacists, they are very challenging to pharmacists and pharmacists don’t have the clinical knowledge to be able to sift through what can often be quite dramatic presentations.*” (GP 1, GP FG1)

#### c. Challenges around anticholinergic medicines

Anticholinergic medicines, which inhibit the neurotransmitter acetylcholine involved in numerous physiological functions, has been associated with adverse outcomes such as cognitive decline and falls, particularly when multiple anticholinergic medicines are used concurrently (termed anticholinergic burden).(15, 16) GPs, clinical pharmacologists, and pharmacists described the importance of reviewing and deprescribing anticholinergic medicines where possible. However, the process of calculating anticholinergic burden (ACB) in frail, older adults is time consuming, primarily due to the absence of automated calculators embedded within the EHR system.

Doctors and pharmacists expressed frustrations around the re-prescribing of anticholinergics after deprescribing them. They attributed the persistence of high ACB to limitations in prescribing guidelines and a scarcity of alternative options to replace anticholinergic drugs. These challenges highlighted the need for tools within EHR systems to facilitate efficient assessment of ACB, alongside a broader exploration of prescribing guidelines and alternatives to enhance deprescribing practices.

> “*One of the things that I often see in general practice is that there’s lots of anticholinergics, usually amitriptyline because it’s kind of given out for other reasons for what it’s licensed for. So, sleep is probably the most common thing that I see it used for, or avoiding long term opioids in chronic arthritic pain, and often that’s because we have other options for them but we’re not allowed to prescribe them. So, melatonin is probably the most common thing that we could put them on which has a lot better safety profile, but we are just completely discouraged from prescribing it. And likewise access to other interventions that would help arthritic pain rather than putting them on NSAIDS which obviously carry risk or opioids which aren’t overly effective outside the acute pain window. It’s often the lack of other stuff that raises all of the anticholinergic burden.*” (GP 1, GP FG1)

Participants welcomed any digital tool that could streamline routine work processes, including information retrieval, automated dose calculations, and assessing the risk of developing diseases to optimise medicines during a SMR in a patient-centred manner, with the goal of enhancing efficiency in the medication optimisation process.

## Discussion

Medication reviews by HCPs can take significant preparation, and are time consuming, primarily due to the need to gather and understand patient information and to develop an understanding of a patient’s medical history and social circumstances. In addition, currently, there is no easy way to identify from the EHR which patients are at greatest risk of medication-related harm and those most likely to benefit from an SMR. The EHR systems used in primary care contain enormous volumes of information which becomes particularly challenging and time-consuming to navigate for complex individuals living with multiple long-term conditions and taking many medications. The way that information is organised in the system leads to a large proportion of time spent linking medications to their original indication and examining the patient journey. This time could be better spent discussing shared decisions with the patient. The EHR has not evolved in line with increasing patient complexity. The findings of this report emphasise the need for enhanced functionalities in EHRs to support effective medication management in the context of deprescribing discussions where a nuanced understanding of a patient’s medication history is crucial.

Our study has highlighted the challenges facing those undertaking SMRs in more socioeconomically disadvantaged areas, where people experience multimorbidity (and co-existent polypharmacy) 10-15 years earlier than their affluent peers.(17, 18) These populations have complex healthcare needs at a younger age, the care of which falls to the already over-stretched GPs. Areas with greater socioeconomic disadvantage often have lower health literacy, resulting from a combination of lower educational attainment, economic barriers like the need to prioritise food and heating over health seeking, and psychosocial stressors affecting decision-making relating to health.(19, 20) Health literacy applies not only to the patient but to the clinician who may also be unaware of the psychosocioeconomic situation of the patient, leading to a communication gap when discussing the risk and benefit of medicines to reach a shared-decision. Accordingly, complex conversations involving numeracy calculations of risk may take longer and require repetition, but may also be of less priority for the patient and/or carer than other more immediate life concerns.

As preparation time is repeatedly cited as a barrier to effective SMR, a potential solution that would support SMRs in those with lower health literacy should include any digital intervention that saves on preparation time. This would enable more time for the clinician to engage with the patient and discuss complexities around risk and benefit, which would go some way to addressing the existing health disparity that affects those experiencing socioeconomic disadvantage. For HCPs working in areas of socioeconomic deprivation, lack of HCP capacity alongside patients declining SMR invitations were cited as barriers to undertaking SMRs. Moreover, HCPs described the usefulness of a system to identify availability of different health services in surrounding areas (e.g. weight management service) and the need to adjust how SMRs are conducted if the patient is from a minority ethnic background. This is important as there is research showing that older people with frailty who are of an ethnic minority background and living in deprived areas are more exposed to polypharmacy and potentially avoidable adverse effects.(17, 18) A recent study co-produced SMR resources to empower patients in their healthcare and support them in making the most out of their SMR. This included producing resources in a number of different languages including audio recorded resources for patients with visual impairment.(21, 22) Embedded links to resources for HCPs to provide to patients before and/or after an SMR can be one potential way to utilise digital health and empower patients to reduce inequity in access to healthcare.

Our study also highlighted medication-related challenges such as difficulties managing mental health, specialist and anticholinergic medications. HCPs reported that a lack of alternatives to medication for symptom management hampered their ability to optimise some of the more potentially harmful medication classes such as opioids, anti-depressants, anticholinergics and gabapentinoids. Non-pharmacological alternatives, where appropriate, such as counselling need to be readily and equitably accessible for this approach to be considered a reliable option.(23) Mental health medication management stood out as a consistent challenge. HCPs in our study, regardless of their professional background, expressed difficulty in monitoring and adjusting psychiatric drugs. There was also a general lack of confidence and skill when it came to monitoring and adjusting antipsychotic medications, with the measurement of success in managing these medications being ambiguous. This is consistent with previous studies that note GPs lack of confidence in managing patients with serious mental health illness.(24, 25) One recent study reported that less than half of GP trainees in England and Wales have trained in a mental health setting between 2013 and 2015.(26) In addition to the need for HCPs in primary care to become trained to address issues related to psychiatric medicines, EHRs must include basic information about the indication for the prescribed psychiatric medicine and the appropriate mental health team contact details for GPs to be able to address these issues. This would have the potential to enable multidisciplinary coordination of care with mental health patients.

Another challenging group of medicines was those with anticholinergic effects. This drug class was also a recurring issue among GPs, clinical pharmacologists, and pharmacists. HCPs found it time consuming to calculate the ACB in frail, older adults. Although there are several ACB scales available that have been developed and validated, participants stated that automated calculators to calculate ACB are not easily accessible or embedded into EHRs. In addition, there is considerable variability between anticholinergic scales making it difficult to ascertain which scale to use to calculate ACB.(27) As such, taking the time out to include every medicine a patient is taking to calculate their ACB is time consuming, reducing opportunities for potential deprescribing discussions with patients.

### Limitations

This study was conducted in the UK, which provides universal access to healthcare. However, findings from our study are to a certain degree globally generic, including the need for HCP coordination (28, 29) or deprescribing challenges.(30, 31) This study is part of a larger qualitative study examining both barriers to SMRs and potential digital solutions, including AI-assisted approaches. As such, the HCP participants likely included a number of clinicians with a particular interest in digital-driven solutions in healthcare. We sought to include a wide variety of HCPs from different practice backgrounds in order to mitigate this.

## Conclusions

There are few useful digital tools that can identify patients that would benefit most from an SMR or monitor the effects of medication optimisation when medicines are altered. Our findings showed that significant time is needed to prepare and conduct a SMR, with complex patients sometimes needing multiple appointments to enable a comprehensive review. The DynAIRx project will use findings from this study to address the barriers of conducting an SMR by producing dashboards and visualisations to summarise the patient’s medical journey; develop digital tools to prioritise patients that would benefit most from an SMR; and identify optimal interventions for specific multimorbidity and polypharmacy patient groups.

## Data Availability

All relevant data are within the manuscript and its Supporting Information files

## Acknowledgements

DynAIRx has been funded by the National Institute for Health and Care Research (NIHR) Artificial Intelligence for Multiple Long-Term Conditions (AIM) call (NIHR 203986). MG is partly funded by the NIHR Applied Research Collaboration North West Coast (ARC NWC). This research is supported by the NIHR ARC NWC. The views expressed in this publication are those of the author(s) and not necessarily those of the NIHR or the Department of Health and Social Care.

## References

1. NHS. Network Contract Directed Enhanced Service. Structured medication reviews and medicines optimisation: guidance. In: NHS, editor. 2020.

2. NHS. Network Contract Directed Enhanced Service. Contract specification 2021/22 - PCN Requirements and Entitlements. 2021.

3. Joseph RM, Knaggs RD, Coupland CAC, Taylor A, Vinogradova Y, Butler D, et al. Frequency and impact of medication reviews for people aged 65 years or above in UK primary care: an observational study using electronic health records. BMC Geriatrics. 2023;23(1):435.

4. Madden M, Mills T, Atkin K, Stewart D, McCambridge J. Early implementation of the structured medication review in England: a qualitative study. British Journal of General Practice. 2022;72(722):e641–e8.

5. Baqir W, Hughes J, Jones T, Barrett S, Desai N, Copeland R, et al. Impact of medication review, within a shared decision-making framework, on deprescribing in people living in care homes. Eur J Hosp Pharm. 2017;24(1):30–3.

6. NICE. Multimorbidity and polypharmacy 2017 [Available from: https://www.nice.org.uk/advice/ktt18.

7. Walker LE, Abuzour AS, Bollegala D, Clegg A, Gabbay M, Griffiths A, et al. The DynAIRx Project Protocol: Artificial Intelligence for dynamic prescribing optimisation and care integration in multimorbidity. Journal of Multimorbidity and Comorbidity. 2022;12:26335565221145493.

8. Heckathorn DD. SNOWBALL VERSUS RESPONDENT-DRIVEN SAMPLING. Sociol Methodol. 2011;41(1):355–66.

9. Clarke V, Braun V. Thematic analysis. The journal of positive psychology. 2017;12(3):297–8.

10. Nowell LS, Norris JM, White DE, Moules NJ. Thematic Analysis:Striving to Meet the Trustworthiness Criteria. International Journal of Qualitative Methods. 2017;16(1):1609406917733847.

11. Ardens Healthcare Informatics. The leaders in providing EMIS Web & SystmOne templates & resources [Available from: https://www.ardens.org.uk/.

12. PARM Diabetes. A health management tool for people with diabetes co-created by Lilly and NHS Devon CCG [Available from: https://parmdiabetes.co.uk/.

13. NHS. Network Contract Directed Enhanced Service. Investment and Impact Fund 2020/21: guidance. 2020.

14. NHS Digital. SNOMED CT 2023 [Available from: https://digital.nhs.uk/services/terminology-and-classifications/snomed-ct.

15. Salahudeen MS, Duffull SB, Nishtala PS. Anticholinergic burden quantified by anticholinergic risk scales and adverse outcomes in older people: a systematic review. BMC Geriatrics. 2015;15(1):31.

16. Mehdizadeh D, Hale M, Todd O, Zaman H, Marques I, Petty D, et al. Associations Between Anticholinergic Medication Exposure and Adverse Health Outcomes in Older People with Frailty: A Systematic Review and Meta-analysis. Drugs - Real World Outcomes. 2021;8(4):431–58.

17. Bennett JE, Pearson-Stuttard J, Kontis V, Capewell S, Wolfe I, Ezzati M. Contributions of diseases and injuries to widening life expectancy inequalities in England from 2001 to 2016: a population-based analysis of vital registration data. The Lancet Public Health. 2018;3(12):e586–e97.

18. Bosworth B. Increasing Disparities in Mortality by Socioeconomic Status. Annual Review of Public Health. 2018;39(1):237–51.

19. Chinn D. Critical health literacy: A review and critical analysis. Social Science & Medicine. 2011;73(1):60–7.

20. Abel T, Benkert R. Critical health literacy: reflection and action for health. Health Promotion International. 2022;37(4).

21. Silcock J, Marques I, Olaniyan J, Raynor DK, Baxter H, Gray N, et al. Co-designing an intervention to improve the process of deprescribing for older people living with frailty in the United Kingdom. Health Expect. 2023;26(1):399–408.

22. Health Innovation Network. Resources to support patients having a Structured Medication Review [Available from: https://thehealthinnovationnetwork.co.uk/programmes/medicines/polypharmacy/patient-information/.

23. Hyde J, Calnan M, Prior L, Lewis G, Kessler D, Sharp D. A qualitative study exploring how GPs decide to prescribe antidepressants. Br J Gen Pract. 2005;55(519):755–62.

24. Nash A, Kingstone T, Farooq S, Tunmore J, Chew-Graham CA. Switching antipsychotics to support the physical health of people with severe mental illness: a qualitative study of healthcare professionals’ perspectives. BMJ Open. 2021;11(2):e042497.

25. Woodall AA, Abuzour AS, Wilson SA, Mair FS, Buchan I, Sheard SB, et al. Management of Antipsychotics in Primary Care: Insights from Healthcare Professionals and Policy Makers in the UK. medRxiv. 2023:2023.11.13.23298487.

26. MIND. Better equipped, better care. Improving mental health training for GPs and practice nurses 2016 [cited 2023 December 11]. Available from: https://www.mind.org.uk/media-a/4501/find-the-words-report-better-equipped-better-care.pdf.

27. Hanlon P, Quinn TJ, Gallacher KI, Myint PK, Jani BD, Nicholl BI, et al. Assessing Risks of Polypharmacy Involving Medications With Anticholinergic Properties. Ann Fam Med. 2020;18(2):148–55.

28. Karam M, Chouinard MC, Poitras ME, Couturier Y, Vedel I, Grgurevic N, et al. Nursing Care Coordination for Patients with Complex Needs in Primary Healthcare: A Scoping Review. Int J Integr Care. 2021;21(1):16.

29. Persson MH, Søndergaard J, Mogensen CB, Skjøt-Arkil H, Andersen PT. Healthcare professionals’ experiences and attitudes to care coordination across health sectors: an interview study. BMC Geriatr. 2022;22(1):509.

30. Ailabouni NJ, Nishtala PS, Mangin D, Tordoff JM. Challenges and Enablers of Deprescribing: A General Practitioner Perspective. PLoS One. 2016;11(4):e0151066.

31. Cullinan S, Raae Hansen C, Byrne S, O’Mahony D, Kearney P, Sahm L. Challenges of deprescribing in the multimorbid patient. Eur J Hosp Pharm. 2017;24(1):43–6.

