## Supplementary material for "A qualitative exploration of barriers to efficient and effective Structured Medication Reviews in Primary Care: Findings from the DynAIRx study": S1 Appendix

**SUPPORTING INFORMATION 1 APPENDIX**

**Acronyms**

AI : Artificial Intelligence

ANP : Advanced Nurse Practitioner

DES : Directed Enhanced Service contract

EHR : Electronic Health Record

GP : General Practitioner

HCP : Health Care Professional

NHS : National Health Service

NICE : National Institute for Health and Care Excellence

PCN : Primary Care Network

SMRs : Structured Medication Reviews

UK : United Kingdom

**Topic guides**

**Healthcare Practitioner Focus Group Topic Guide**

The purpose of these focus groups with key stakeholders is to draw out some common themes on challenges to undertaking structured medicines reviews, requirements to do them well and opportunities to improve.

Background: I will give a short (5 minute) overview about the Overprescribing review and structured medication reviews to introduce the topic. We will play the 2 minute DynAIRx animation [https://youtu.be/cKKTYTkWIEY](https://gbr01.safelinks.protection.outlook.com/?url=https%3A%2F%2Fyoutu.be%2FcKKTYTkWIEY&data=05%7C01%7Cwasim.baqir%40nhs.net%7C3267ce3e24f74b106f0008dac0a582bd%7C37c354b285b047f5b22207b48d774ee3%7C0%7C0%7C638034114341973645%7CUnknown%7CTWFpbGZsb3d8eyJWIjoiMC4wLjAwMDAiLCJQIjoiV2luMzIiLCJBTiI6Ik1haWwiLCJXVCI6Mn0%3D%7C3000%7C%7C%7C&sdata=GQS4uE3uRltmPrUen1fI61yaDDcqDIUbWtZjoWC%2FAFg%3D&reserved=0)

I will introduce the potential opportunity that AI presents to augment SMRs.

**Topic 1: How are Structured Medication Reviews currently being undertaken, by whom and where, and how long do they take?**

How long are the appointments allocated versus how long do they actually take per patient? Who does them? What is the process for escalation?

Probe: What are the first steps? What resources do you need to when conducting a SMR (letters, blood results)? What information would aid the medicines optimisation process in a SMR? Is there usually any information missing/you are unable to retrieve that would assist you in the SMR process? (e.g. discharge letters and poor information flow from secondary to primary care)

Is there a patient prioritisation process when it comes to inviting patients for a SMR? How do you decide in your setting who should be prioritized? Is it based on number of drugs? Complexity? Prescribing indicators? Frailty? Who does the prioritization? Can you give examples of the type of patients you usually prioritise? Why do you choose to prioritise them?

**Topic 3: What do participants consider the top priority target medication challenges relating to key multimorbidity groups (older people with frailty; co-existing physical and mental health problems; complex multimorbidity and potentially problematic polypharmacy)?**

Probe: What kind of medication-related challenges do you experience when preparing for/conducting a SMR? What are the risky medication combinations that present the biggest challenges when optimising medication? What are the hardest medications to withdraw? How many (relevant) diagnoses do they tend to have? How many different medications are they on? Typical/min/max.

**Topic 4: What kind of digital tools do they think would be most helpful for assisting efficient SMRs?**

Describe what a digital tool is and what it might look like so that they are able to answer the question.

What tools are used currently – why – what are the benefits, drawbacks?

To prompt this I could show examples of currently available digital tools and some idea of how long they take to implement

**Topic 5: With digital tools in mind, what kind of data would prescribers and practitioners involved in reviewing medications need to undertake effective Structured Medication Reviews efficiently?**

Probe: What do they agree with? What do they disagree with? Specifically focusing on the notion of AI to aid digital summarising of patient trajectories of risk – what are perceptions/ideas about the role of an AI-intervention?

**Topic 6: What are likely barriers/facilitators to uptake and utilisation of AI(-augmented) tools?**

What would be a sign to them that the intervention is useful?

Probe: What does success look like (number of reviews undertaken? most high-risk people prioritized?)

What outcomes would they like to see to show it was worth continuing to keep using it? What would put them off? How would participants feel about being involved in trying out the intervention during the development phase, in order to feedback and improve it?

**Any further areas of discussion:**

- Do any of the participants have anything further to add from what we have discussed today?

**Give thanks for participating.**

**Patient Focus Group Topic Guide**

The contents of this focus group guide will be informed by the findings of the ongoing one-on-one scoping interviews with key stakeholders so may be subject to some changes in content.

**Background:** I will give a short (10 minute) general presentation about the problems facing health care professionals who are trying to deliver structured medicines reviews to patients with multimorbidity and/or polypharmacy, and the themes that have emerged from scoping interviews with key stakeholders about the possible barriers/facilitators to effective SMRs

**The following issues will then be addressed/explored:**

**Topic 1: How are Structured Medication Reviews currently being undertaken, by whom and where, and how long do they take?**

What are the consultations like? How long are the appointments allocated versus how long do they actually take? Who does them? Do you have enough time to discuss and ask questions about your medications?

Probe: Do you get a chance to discuss all of your medications, or are only some discussed? How does the health care professional interact and explain benefits and risks of your medicines with you?

**Topic 2: What data do prescribers and practitioners involved in reviewing medications need to undertake effective Structured Medication Reviews efficiently?**

What do you the patients think about the suggestions and themes that have merged from initial scoping interviews?

Probe: What do they agree with? What do they disagree with? Specifically focusing on the notion of AI to aid digital summarising of patient trajectories of risk – what are perceptions/ideas about the role of an AI-intervention? Would you have confidence in a decision support tool assisting your doctor? Do you think using it would aid or hinder the consultation? Do you think your priorities around your health and medication wishes would be addressed?

**Topic 3: What do participants consider the top priority target medication challenges relating to key multimorbidity groups (older people with frailty; co-existing physical and mental health problems; complex multimorbidity and potentially problematic polypharmacy)?**

Probe: What do you think as a patient your healthcare professional may find difficult in reviewing your medications with you? Are your aims the same as the healthcare professional?

**Topic 4: What kind of digital tools do they think would be most helpful for assisting efficient SMRs?**

What tools are used currently by your healthcare professional during the consultation – why – what are the benefits, drawbacks? Does your healthcare professional using these tools (see examples given) help or hinder the consultation?

To prompt this, I could show examples of currently available deprescribing resources and some idea of how long they take to implement (Drug burden index, BNF interaction checker).

**Topic 5: What are likely barriers/facilitators to uptake and utilisation of AI(-augmented) tools?**

What do you think would affect your healthcare professional using these tools during a medication review with you?

Probe: What would a successful decision support tool deliver for you, the patient?

What outcomes would they like to see to show it was worth continuing to keep using it? What would put them off being in a consultation if the tool was being used? How would patients feel about being involved in trying out the intervention during the development phase with your healthcare professional, in order to feedback and improve it?

**Any further areas of discussion:**

- Do any of the participants have anything further to add from what we have discussed today?

**Give thanks for participating.**
